# Precarious employment and associations with health during COVID-19: a nationally representative survey in Wales, UK

**DOI:** 10.1101/2022.06.07.22275493

**Authors:** Benjamin J Gray, Richard G Kyle, Kate R Isherwood, Ciarán Humphreys, Melda Lois Griffiths, Alisha R Davies

## Abstract

**Background:** The COVID-19 pandemic had an early impact on employment, with the United States (US) and the United Kingdom (UK) experiencing more severe immediate labour market impacts than other Western countries. Emerging evidence from the initial phase of the pandemic highlighted that job losses were experienced more by those holding atypical contracts. Furthermore, it is predicted that this associated unemployment will increase precarious employment arrangements during the COVID-19 pandemic.

In this paper we seek to answer the following research questions:

I. What is the prevalence of precarious employment in Wales and are there differences in employment precariousness by socio-demographic characteristics and self-reported health status?
II. Which domains are the main contributing factors of precarious employment in Wales?
III. Which domains of precarious employment are associated with poorer health?
IV. Haves there been changes in job quality (as reflected by precarious employment domains) during the COVID pandemic (between February 2020 and Winter 2020/2021)?

**Methods:** Data was collected from a national household survey carried out in May/June 2020, with a sample of 1,032 residents in Wales and follow-up responses from 429 individuals collected between November 2020 and January 2021. To examine the associations between experiencing precarious employment or the separate domains of employment precariousness and socio-demographics and health, chi-squared analyses and logistic regression models (multinomial and binary) were used. To determine longitudinal changes in precarious employment experienced by socio-demographic groups and furlough status, McNemar’s test was used. The data is presented as proportion of respondents or adjusted odds ratios (aOR) and 95% confidence intervals following logistic regression.

**Results:** Overall, pre-pandemic, one in four respondents were determined to be in precarious employment (26.5%). A higher proportion of females (28.3%) and those aged 18-29 years (41.0%) were in precarious employment in February 2020. In addition, a greater percentage of individuals who reported poorer health across all self-reported measures were in precarious employment compared to those reporting better health. Worse perceived treatment at work was twice as likely in those who reported a pre-existing condition (aOR 2.45 95% CI [1.33-4.49]), poorer general health (aOR 2.33 95% CI [1.22-4.47]) or low mental wellbeing (aOR 2.81 95% CI [1.34-5.88]) when compared to their healthier counterparts. Those calculated to have high wage precariousness were three times more likely to report low mental wellbeing (aOR 3.12 95% CI [1.54-6.32]). In the subsample, there was an observed increase in the prevalence of precarious employment, with this being attributable to lower affordability of wages and a perceived increase in vulnerability at work. The subgroups that were most impacted by this decrease in job quality were females and the 30-39 years age group.

**Implications:** Improving the vulnerability and wages domains, through the creation and provision of secure, adequately paid job opportunities has the potential to reduce the prevalence of precarious employment in Wales. In turn, these changes would improve the health and wellbeing of the working age population, some of which are already adversely impacted by the COVID-19 pandemic.

## Background

The COVID-19 pandemic had an early impact on employment, with the United States (US) and the United Kingdom (UK) experiencing more severe immediate labour market impacts than other Western countries [1,2]. Emerging evidence from the initial phase of the pandemic highlighted that job losses were experienced more by those holding atypical contracts [3]. Furthermore, past global recessions and associated high unemployment rates have disempowered workers [4,5] and resulted in an increase of precarious employment arrangements. Based on these previous events, it has also been predicted that precarious employment will also increase during the COVID-19 pandemic as a result of the forecasted high rates of unemployment [6].

One of the limitations to much of the international literature is the reliance on contract type as a proxy for precarious employment [7,8]. However, precarious employment is a multi-dimensional construct, encompassing more than contract type [7] and it is critical that future research examines precarious employment from this multi-dimensional perspective [8]. There is no single definition of precarious employment, but it is recognised as a multi-dimensional construct encompassing dimensions of: employment insecurity, incorporating both length of contract and perceptions of job insecurity; individualised bargaining; relations between workers and employers; low wages and economic deprivation; limited workplace rights and social protection; and powerlessness to exercise legally granted workplace rights [8,9].

The Employment Precariousness Scale (EPRES) has been developed to assess the multi-dimensional construct of precarious employment in its calculation. The EPRES was developed in Spain [9], adapted for use in Sweden [10] and measures six domains of precarious employment. To date, the EPRES has not been used previously in the UK and therefore the prevalence of precarious employment is under-researched and the impacts on health not fully understood. Better understanding of which components of precarious employment are more strongly associated with poor health can help to direct action by policy makers and employers themselves, to address the key factors of precarious employment with the greatest potential to address underlying health. This article addresses the following research questions using data from a nationally representative survey in Wales, UK:

I. What is the prevalence of precarious employment in Wales and are there differences in employment precariousness by socio-demographic characteristics and self-reported health status?
II. Which domains are the main contributing factors of precarious employment in Wales?
III. Which domains of precarious employment are associated with poorer health?
IV. Have there been changes in job quality (as reflected by precarious employment domains) during the COVID pandemic (between February 2020 and Winter 2020/2021)?

## Methods

### Data Source

The cross-sectional data presented here was collected in May/June 2020 as part of the *COVID-19, Employment and Health in Wales* study, a national household survey. The initial recruitment was a postal invitation and follow-up letter 2 weeks later encouraging online completion of a questionnaire (“push to web approach”) delivered to a stratified random sample of 20,000 households (response rate 6.9%). Each selected household was sent a survey pack containing an invitation letter and participant information sheet. The invitation asked the eligible member of the household with the next birthday to participate in the survey. It included instructions on how to access the online questionnaire, by entering a unique reference number provided in the letter. Full details of the initial study recruitment are discussed extensively elsewhere [3]. The original survey also asked for the individual to consent to opt-in to a future follow-up study. Of the 1,379 respondents, 1,084 consented (78.6%) to be contacted and optionally provided their names and email addresses. The follow-up data collection phase was from November 2020 to January 2021 (Winter 2020/21). If a valid email address was provided (n=925, 85.3%), individuals were emailed an invitation to take part with two further email remainders to encourage participation. If a valid email address was not provided (n=159, 14.7%), individuals were sent a postal invitation and one reminder invitation. In total, 626 individuals completed the follow-up online questionnaire (58% response rate).

### Eligibility Criteria

Respondents included in the baseline sample were in ‘paid employment’, i.e., not self-employed in February 2020 and had to provide complete data for all five included domains of precarious employment (n=1,032; 74.8% of total respondents). Longitudinal analysis was performed on the individuals who completed the follow-up questionnaire, were in ‘paid employment’ in February 2020 and Winter 2020/21 and completed all five domains of precarious employment (n=429; 41.6% of baseline sample). Participants who completed the follow-up questionnaire and indicated that they has been furloughed at any point during the pandemic were also included in the longitudinal analysis.

### Questionnaire Measures

Explanatory variables included; socio-demographics (gender, age group, and deprivation quintile assigned based on postcode of residence using the Welsh Index of Multiple Deprivation (WIMD [11])); individual self-reported health status included general health and pre-existing health conditions (assessed using validated questions from the National Survey for Wales [12]) and mental wellbeing (assessed using the Short Version of the Warwick Edinburgh Mental Wellbeing Score [13]). In line with established research practice, we determined low mental wellbeing as 1 standard deviation below the mean score [14]. Precarious employment was determined using the calculation from the EPRES [9]. The questions that form each of the five domains are shown in Table 1, some of which have been amended for a UK context. For comparative purposes, the original EPRES questionnaire is provided as supplementary material (Supplementary Table S1). For each domain, the scores of the responses were totalled, divided by the highest possible score and then multiplied by four. This domain score would fall between 0-0.999 (no/low precariousness), 1-1.999 (moderate precariousness) and 2-4 (high to very high precariousness). For the overall score, the total sum of the domains were added together and divided by five and this value would again fall between the ranges and associated levels of precariousness outlined previously [9].

**Table 1.**
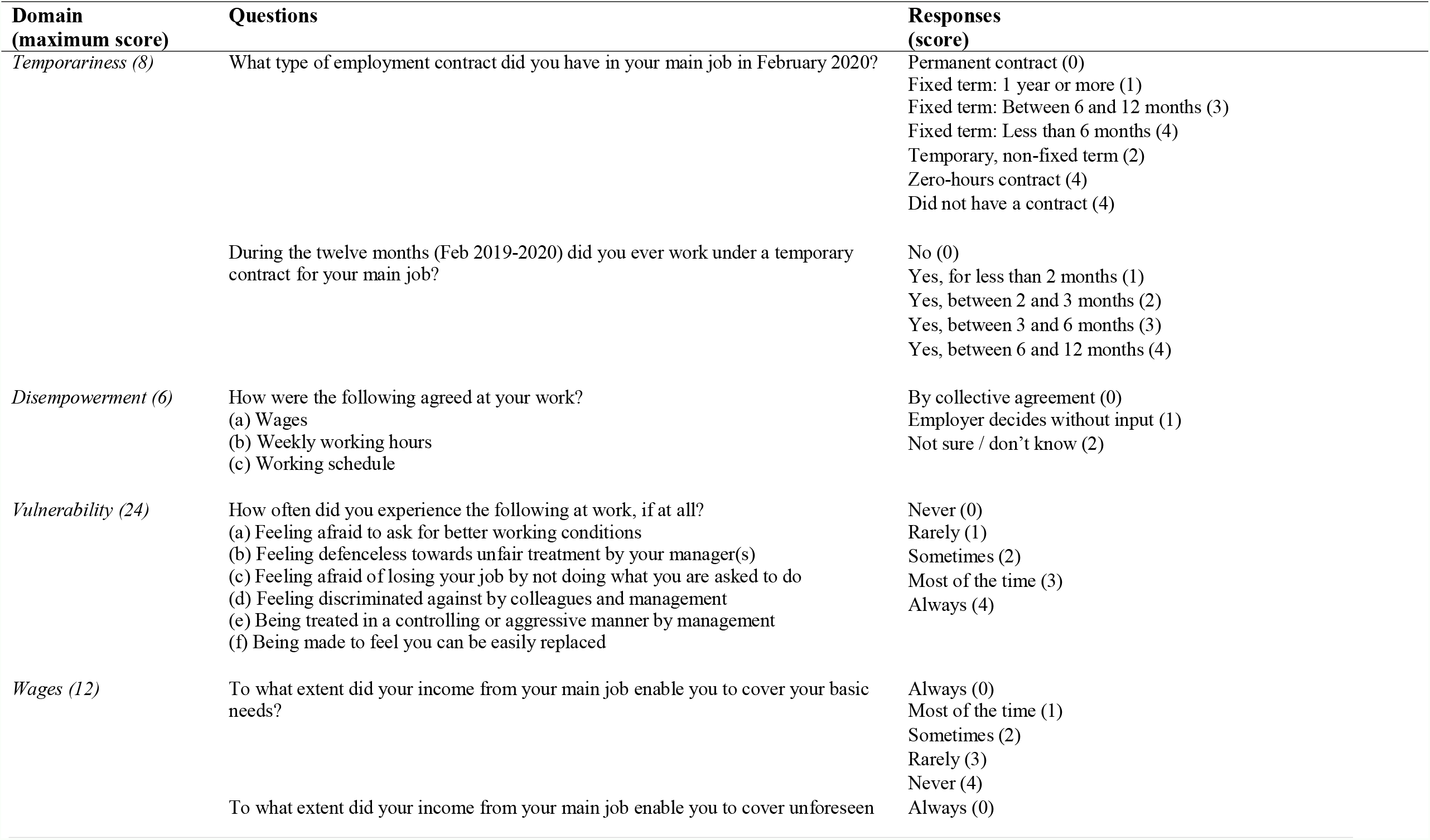
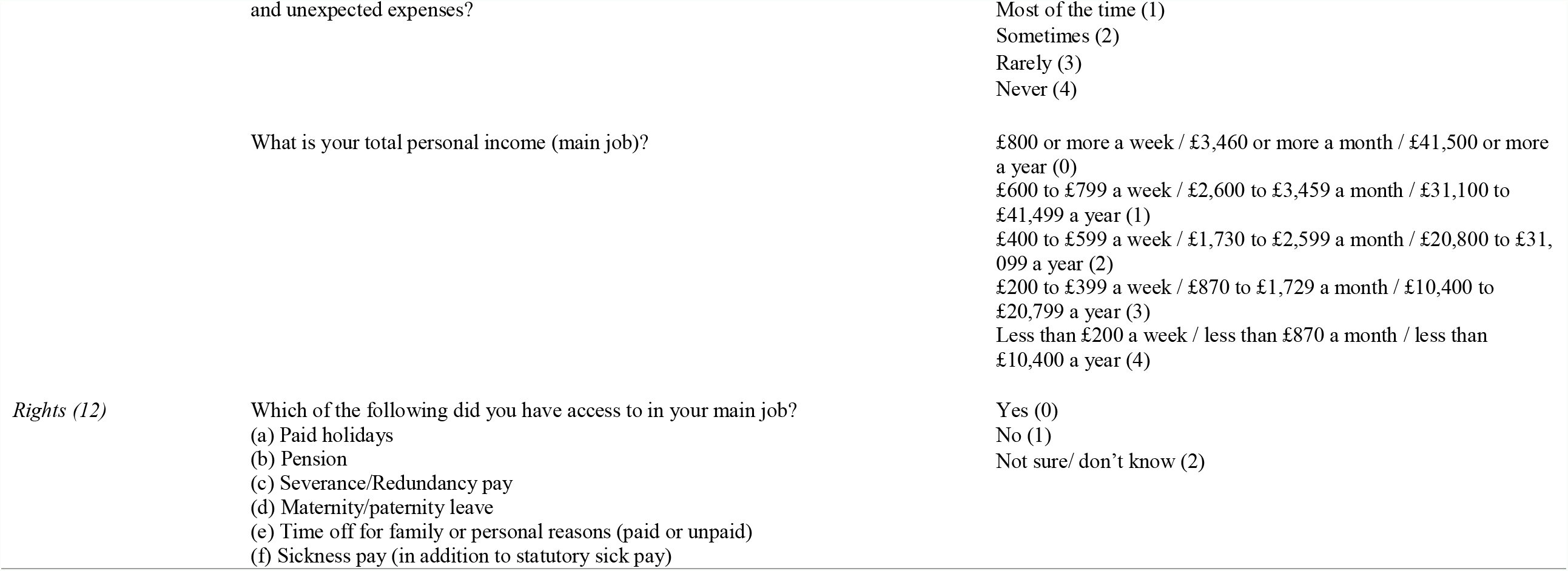
Questions and responses contained within the five domains of precarious employment included in this study.

### Statistical Analysis

To examine the associations between experiencing precarious employment or the separate domains of employment precariousness and socio-demographics and health, chi-squared analyses and logistic regression models (multinomial and binary) were used. To determine longitudinal changes in precarious employment experienced by socio-demographic groups and furlough status, McNemar’s test was used. The data is presented as proportion of respondents or adjusted odds ratios and 95% confidence intervals following logistic regression. P value was set at <0.05 for statistical significance.

## Results

### Sample Characteristics

A higher proportion of respondents were female, the age range with the greatest number of responses were 40-59 years and the responses across deprivation quintile were well distributed (Table 2). Although generally representative, females and older age groups are over-represented, while the youngest age group is under-represented, when comparing our sample to the Welsh “working age” population. There were no statistically significant differences between the sociodemographic characteristics of the baseline sample (n=1,032) and the longitudinal sub-sample (n=429; Table 2).

**Table 2.** Socio-demographic characteristics of the baseline and longitudinal analysis samples.

|  | Precarious<br>Employment Baseline<br>Sample<br>(n=1,032) | Precarious<br>Longitudinal Analysis<br>Sample<br>(n=429) | Welsh 'working age'<br>Population<br>mid-year 2018<br>(n=1,856,853) |
| --- | --- | --- | --- |
| *Gender* |  |  |  |
| Male | 388 (37.6%) | 146 (34.0%) | 924,020 (49.8%) |
| Female | 636 (61.6%) | 282 (65.7%) | 932,833 (50.2%) |
| *Age Group* |  |  |  |
| 18-29 Years | 134 (13.0%) | 39 (9.1%) | 485,909 (26.2%) |
| 30-39 Years | 217 (21.0%) | 91 (21.2%) | 371,851 (20.0%) |
| 40-49 Years | 262 (25.4%) | 119 (27.7%) | 375,526 (20.2%) |
| 50-59 Years | 295 (28.9%) | 125 (29.1%) | 433,915 (23.4%) |
| 60-64 Years | 112 (11.0%) | 54 (12.6%) | 189,652 (10.2%) |
| *Deprivation Quintile* |  |  |  |
| WIMD 1 | 195 (18.9%) | 74 (17.2%) | 371,014 (20.0%) |
| WIMD 2 | 253 (24.5%) | 109 (25.4%) | 370,637 (20.0%) |
| WIMD 3 | 166 (16.1%) | 70 (16.3%) | 384,927 (20.7%) |
| WIMD 4 | 187 (18.1%) | 79 (18.4%) | 370,242 (19.9%) |
| WIMD 5 | 231 (22.4%) | 97 (22.6%) | 360,033 (19.4%) |

### Research Question I. What is the prevalence of precarious employment in Wales and are there differences in employment precariousness by socio-demographic characteristics and self-reported health status?

Overall, pre-COVID-19 pandemic (February 2020), one in four respondents were calculated to be in employment with a moderate or higher level of precarity (26.5%). A higher proportion of females compared to men were in precarious roles (28.3% compared to 22.7%, P=0.047), there were differences in prevalence across age groups, with the highest prevalence (41.0%) reported in the youngest age group (18-29 years). No statistically significant differences were observed across deprivation quintiles, however the highest prevalence (32.3%) was observed in the most deprived quintile (Table 3). Considering self-reported health, a higher proportion of those individuals who reported poorer health were in precarious employment. This observation was consistent across pre-existing health condition (31.9% compared to 22.4%; P=0.001), ‘not good’ general health (35.1% compared to 24.0%, P=0.001) and low mental wellbeing (47.5% compared to 24.3%, P<0.001; Table 3).Differences were also evident by contract type with a higher proportion of people holding non-permanent contracts (Atypical: 90.0%; Fixed-Term: 64.3%;) calculated to be in precarious employment, however, one in five (20.7%) permanent jobs were calculated to be precarious (Table 3).

**Table 3.**
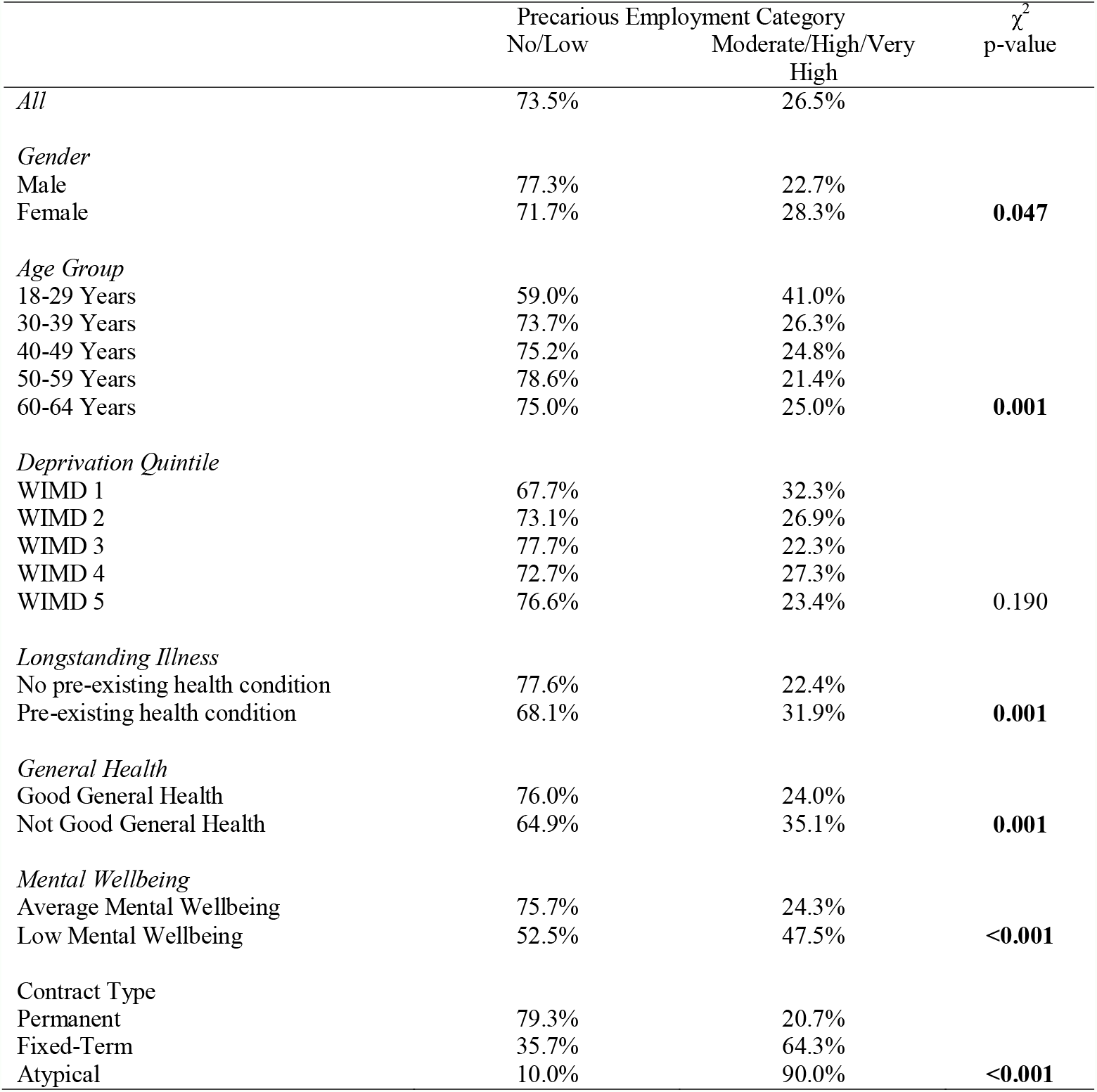
Prevalence of precarious employment in Wales, overall and subgroup analysis

After adjusting for socio-demographic factors and self-reported health, independent predictors of those in precarious employment were females compared to males (aOR 1.42 [95% CI 1.02-1.96]), being of a younger age (18-29 years (aOR 2.26 [95% CI 1.39-3.69]), reporting a pre-existing health condition (aOR 1.44 [95% CI 1.03-2.01]) or low mental wellbeing (aOR 2.33 [95% CI 1.41-3.86]) compared to their healthier counterparts (Table 4).

**Table 4.** Associations of experiencing precarious employment by socio-economic and self-reported health status.

|  | Precarious Employment MLR |
| --- | --- |
|  | No/Low vs.<br>Moderate/High/Very High |
| *Gender* |  |
| Male | Reference |
| Female | **1.42 [1.02-1.96]** |
| *Age Group* |  |
| 18-29 Years | **2.26 [1.39-3.69]** |
| 30-39 Years | 1.08 [0.69-1.70] |
| 40-49 Years | Reference |
| 50-59 Years | 0.88 [0.57-1.35] |
| 60-64 Years | 1.00 [0.57-1.76] |
| *Deprivation Quintile* |  |
| WIMD 1 | 1.38 [0.86-2.22] |
| WIMD 2 | 1.05 [0.66-1.67] |
| WIMD 3 | 0.88 [0.52-1.48] |
| WIMD 4 | 1.27 [0.78-2.07] |
| WIMD 5 | Reference |
| *Longstanding Illness* |  |
| No pre-existing health condition | Reference |
| Pre-existing health condition | **1.44 [1.03-2.01]** |
| *General Health* |  |
| Good General Health | Reference |
| Not Good General Health | 1.43 [0.96-2.14] |
| *Mental Wellbeing* |  |
| Average Mental Wellbeing | Reference |
| Low Mental Wellbeing | **2.33 [1.41-3.86]** |

Adjusted odds ratios (gender, age group, deprivation quintile, longstanding illness, general health and mental wellbeing);

### Research Question II. Which domains are the main contributing factors of precarious employment in Wales?

In our sample, 84.8% of individuals reported at least one domain of precarious employment at a moderate level of precariousness or higher. The most prevalent domains of precarious employment in Wales were found to be *wages* (over 60% either moderate precariousness or higher) and *disempowerment* (over 50% either moderate precariousness or higher). Feelings of *vulnerability at work* were reported in 25% of respondents, whereas 30% of respondents reported a lack of access to employment rights (Figure 1). Only 10% of respondents reported moderate or high precariousness in relation to temporariness (of contract), yet when precariousness is considered as a broader construct i.e. across five domains, over 25% of respondents in paid employment were in roles which were of moderate or higher precariousness (Tables 3 and 4).

**Figure 1.**
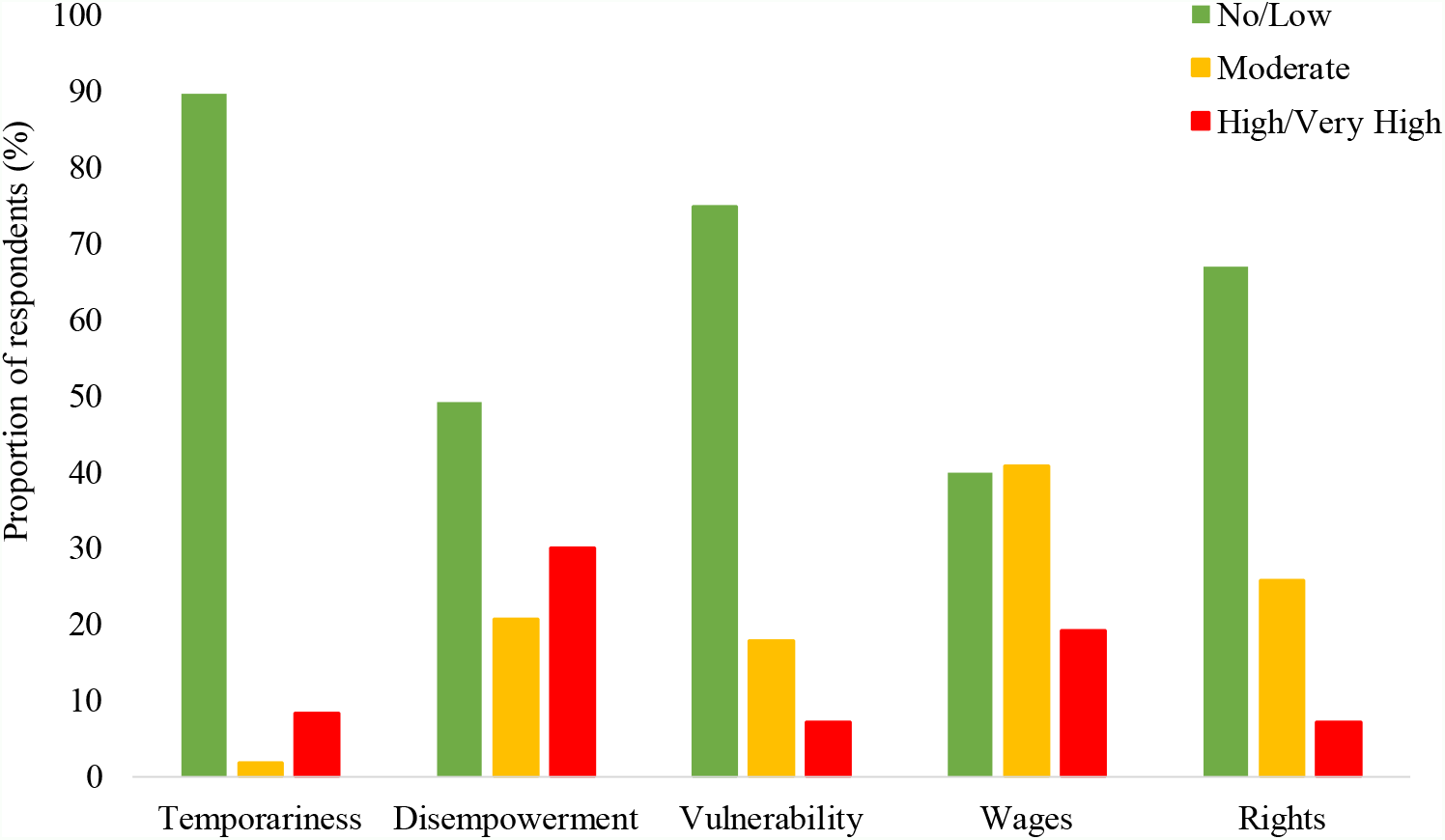
Overall prevalence of the separate precarious employment domains in Wales

Some differences were observed across socio-demographics when experiencing high to very high precariousness (Figure 2). A higher proportion of males experienced high disempowerment precariousness, whilst a greater proportion of females reported wages precariousness (Figure 2A). Across four of the five domains (temporariness, disempowerment, vulnerability and rights) the prevalence of high precariousness was greatest amongst the youngest age group (18-29 years; Figure 2B). Those from the most deprived areas reported the greatest proportions who experienced high precariousness in relation to less rights and low wages (Figure 2C).

**Figure 2.**
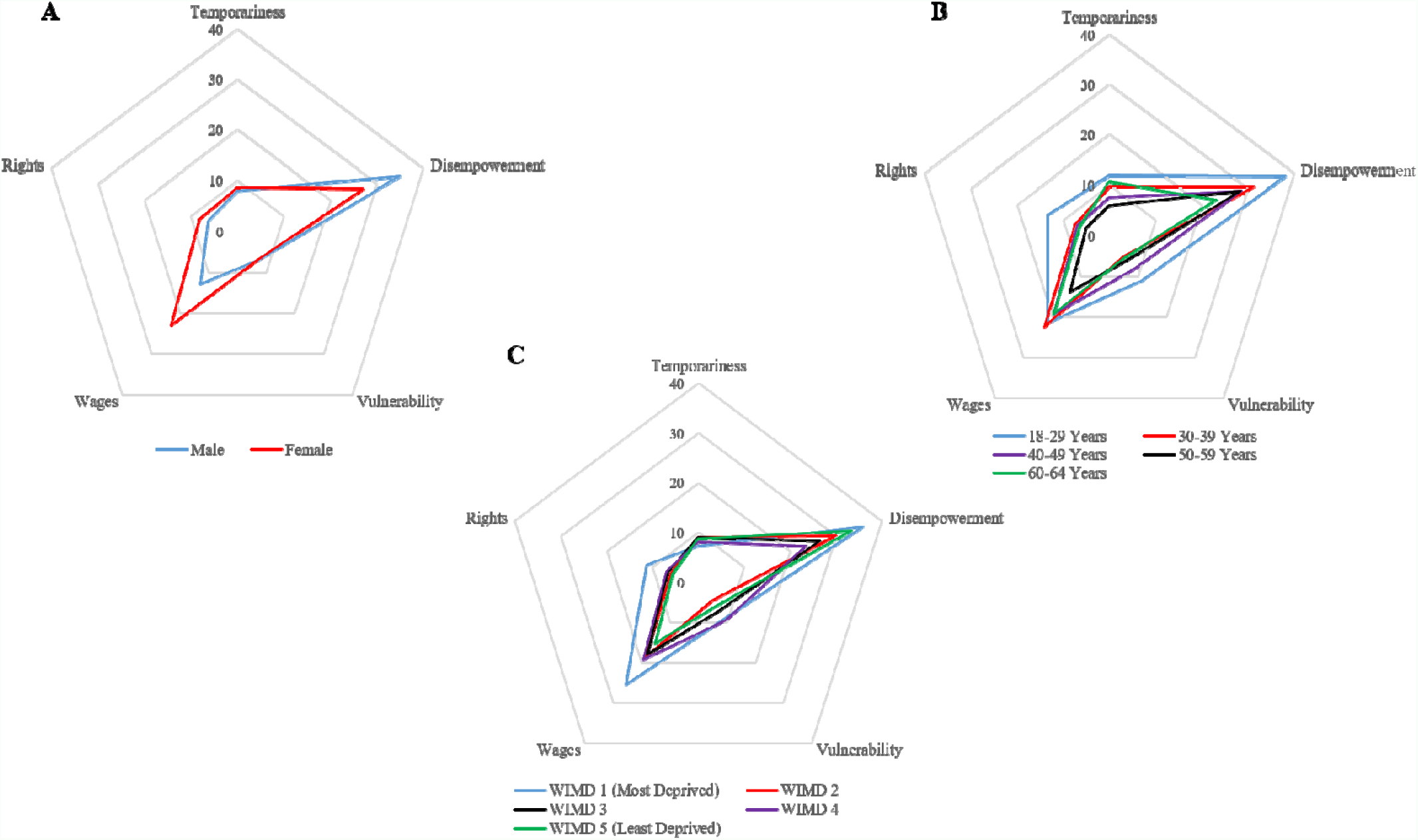
Proportions of socio-demographic groupings calculated at high to very high precariousness across the five domains of precarious employment.

### Research Question II. Which domains of precarious employment are associated with poorer health?

The differences in proportions of high to very high precariousness across the five domains was more clearly observed when comparing health status (Figure 3) than the same comparisons across socio-demographic variables (Figure 2). Proportions of high precariousness are greater amongst four domains (*temporariness, vulnerability, wages, rights*) when considering pre-existing conditions (Figure 3A), two (*vulnerability, wages*) when examining general health status (Figure 3B) and all five with regards to mental health (Figure 3C).

**Figure 3.**
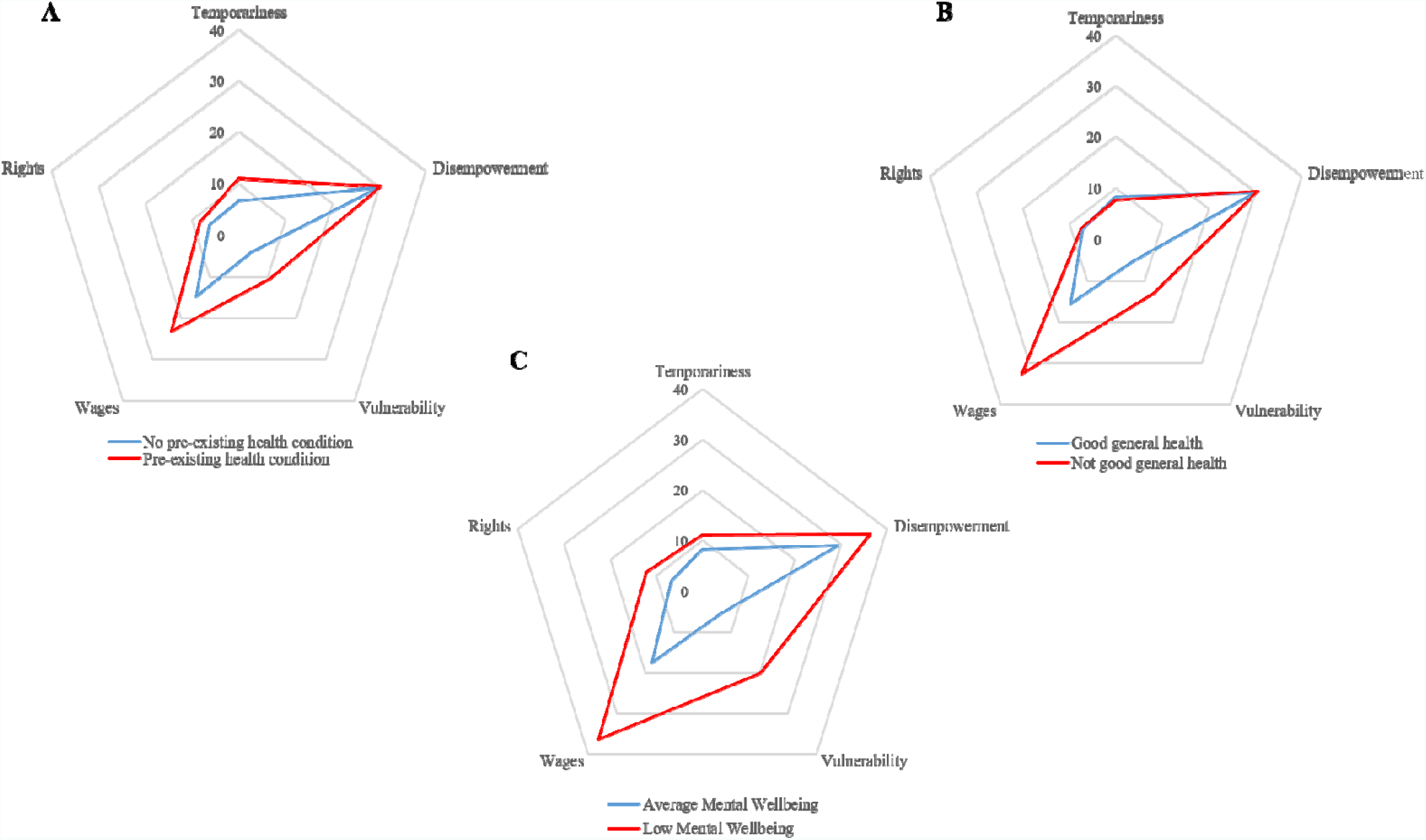
Proportions of self-reported health groupings calculated at high to very high precariousness across the five domains of precarious employment.

The clearest associations between individual domains of precarious employment and poorer health are observed in the *vulnerability* and *wages* domains (Tables 5 and 6). High or very high vulnerability precariousness was over two times more likely in those who reported a pre-existing condition (aOR 2.45 [95% CI 1.33-4.49], not good general health (aOR 2.33 [95% CI 1.22-4.47] or low mental wellbeing (aOR 2.81 [95% CI 1.34-5.88] when compared to their healthier counterparts. The only subgroup that did not observe an association with poorer health and some degree of *wages* precariousness was general health comparisons in the moderate precariousness category. Those calculated to have high *wages* precariousness were three times more likely to report low mental wellbeing (aOR 3.12 [95% CI 1.54-6.32]). Individuals with pre-existing conditions were almost two times more likely to be in high *temporariness* precariousness (aOR 1.90 [95% CI 1.15-3.15]), whereas those reporting not good general health were almost two times more likely to experience a moderate degree of *disempowerment* (aOR 1.93 [95% CI 1.22-3.05]).

**Table 5.** Predictors of experiencing temporariness, disempowerment and vulnerability as determined by multinomial logistic regression.

|  | Temporariness MLR |  | Disempowerment MLR |  | Vulnerability MLR |  |
| --- | --- | --- | --- | --- | --- | --- |
|  | No/Low vs.<br>Moderate | No/Low vs.<br>High/Very High | No/Low vs.<br>Moderate | No/Low vs.<br>High/Very High | No/Low vs.<br>Moderate | No/Low vs.<br>High/Very High |
| *Gender* |  |  |  |  |  |  |
| Male | Reference | Reference | Reference | Reference | Reference | Reference |
| Female | 0.89 [0.32-2.43] | 0.99 [0.61-1.63] | 1.01 [0.71-1.45] | 0.73 [0.54-1.01] | 0.89 [0.62-1.27] | 0.99 [0.55-1.78] |
| *Age Group* |  |  |  |  |  |  |
| 18-29 Years | 2.77 [0.71-10.87] | 1.85 [0.85-4.04] | 1.63 [0.91-2.89] | **1.96 [1.16-3.30]** | 0.72 [0.36-1.42] | 1.45 [0.66-3.19] |
| 30-39 Years | 1.13 [0.27-4.72] | 1.62 [0.81-3.26] | 0.95 [0.58-1.55] | 1.10 [0.71-1.72] | 1.37 [0.84-2.25] | 0.40 [0.16-1.00] |
| 40-49 Years | Reference | Reference | Reference | Reference | Reference | Reference |
| 50-59 Years | 0.90 [0.21-3.72] | 0.95 [0.47-1.94] | 0.72 [0.45-1.15] | 0.90 [0.60-1.37] | 1.40 [0.88-2.23] | 0.60 [0.28-1.28] |
| 60-64 Years | ---- | 1.66 [0.74-3.74] | 0.81 [0.45-1.48] | 0.66 [0.37-1.18] | 0.70 [0.35-1.39] | 0.53 [0.19-1.52] |
| *Deprivation Quintile* |  |  |  |  |  |  |
| WIMD 1 | ---- | 0.78 [0.36-1.71] | 0.95 [0.54-1.65] | 1.00 [0.63-1.59] | 1.12 [0.64-1.94] | 1.80 [0.70-4.60] |
| WIMD 2 | ---- | 1.01 [0.51-2.02] | 1.21 [0.73-2.00] | 0.90 [0.58-1.40] | 1.04 [0.62-1.74] | 0.85 [0.32-2.25] |
| WIMD 3 | ---- | 1.19 [0.57-2.49] | 0.88 [0.50-1.53] | 0.63 [0.38-1.04] | 1.20 [0.68-2.12] | 1.48 [0.55-3.96] |
| WIMD 4 | ---- | 0.92 [0.43-1.97] | 0.90 [0.52-1.54] | **0.60 [0.37-0.98]** | 1.16 [0.69-2.01] | 2.39 [0.97-5.92] |
| WIMD 5 | Reference | Reference | Reference | Reference | Reference | Reference |
| *Longstanding Illness* |  |  |  |  |  |  |
| No pre-existing health condition | Reference | Reference | Reference | Reference | Reference | Reference |
| Pre-existing health condition | 1.02 [0.34-3.08] | **1.90 [1.15-3.15]** | 0.79 [0.54-1.17] | 1.03 [0.73-1.45] | **1.50 [1.03-2.18]** | **2.45 [1.33-4.49]** |
| *General Health* |  |  |  |  |  |  |
| Good General Health | Reference | Reference | Reference | Reference | Reference | Reference |
| Not Good General Health | 1.42 [0.40-5.01] | 0.71 [0.37-1.38] | **1.93 [1.22-3.05]** | 1.09 [0.70-1.69] | **1.83 [1.18-2.84]** | **2.33 [1.22-4.47]** |
| *Mental Wellbeing* |  |  |  |  |  |  |
| Average Mental Wellbeing | Reference | Reference | Reference | Reference | Reference | Reference |
| Low Mental Wellbeing | 1.17 [0.24-5.80] | 1.26 [0.56-2.84] | 1.55 [0.83-2.88] | 1.72 [0.97-3.05] | 1.09 [0.58-2.05] | **2.81 [1.34-5.88]** |

**Table 6.** Predictors of experiencing (low) wages and fewer employment rights as determined by multinomial logistic regression.

|  | Wages MLR |  | Rights MLR |  |
| --- | --- | --- | --- | --- |
|  | No/Low vs.<br>Moderate | No/Low vs.<br>High/Very High | No/Low vs.<br>Moderate | No/Low vs.<br>High/Very High |
| *Gender* |  |  |  |  |
| Male | Reference | Reference | Reference | Reference |
| Female | **2.59 [1.91-3.52]** | **3.39 [2.22-5.16]** | 1.22 [0.89-1.67] | 1.15 [0.66-2.00] |
| *Age Group* |  |  |  |  |
| 18-29 Years | **2.00 [1.18-3.38]** | 1.91 [0.98-3.73] | **2.02 [1.20-3.42]** | **3.11 [1.43-6.75]** |
| 30-39 Years | 0.85 [0.55-1.32] | 1.05 [0.61-1.82] | 1.07 [0.67-1.73] | 1.22 [0.56-2.64] |
| 40-49 Years | Reference | Reference | Reference | Reference |
| 50-59 Years | 0.92 [0.62-1.09] | 0.63 [0.37-1.12] | **1.78 [1.17-2.72]** | 1.02 [0.47-2.23] |
| 60-64 Years | 1.00 [0.58-1.73] | 1.64 [0.85-3.13] | **2.38 [1.40-4.03]** | 1.12 [0.38-3.26] |
| *Deprivation Quintile* |  |  |  |  |
| WIMD 1 | 1.45 [0.90-2.32] | **1.98 [1.09-3.62]** | **1.74 [1.05-2.89]** | 2.02 [0.93-4.41] |
| WIMD 2 | 1.11 [0.72-1.71] | 1.14 [0.64-2.05] | **1.81 [1.13-2.90]** | 1.13 [0.50-2.55] |
| WIMD 3 | 1.45 [0.90-2.33] | 1.28 [0.67-2.44] | **1.88 [1.13-3.14]** | 1.23 [0.51-2.98] |
| WIMD 4 | 1.13 [0.71-1.79] | 1.33 [0.72-2.45] | **2.07 [1.26-3.38]** | 1.06 [0.43-2.62] |
| WIMD 5 | Reference | Reference | Reference | Reference |
| *Longstanding Illness* |  |  |  |  |
| No pre-existing health condition | Reference | Reference | Reference | Reference |
| Pre-existing health condition | **1.70 [1.22-2.38]** | **1.71 [1.11-2.62]** | 1.17 [0.84-1.65] | 1.59 [0.91-2.80] |
| *General Health* |  |  |  |  |
| Good General Health | Reference | Reference | Reference | Reference |
| Not Good General Health | 1.05 [0.68-1.63] | **2.16 [1.31-3.58]** | 1.17 [0.77-1.76] | 0.89 [0.43-1.82] |
| *Mental Wellbeing* |  |  |  |  |
| Average Mental Wellbeing | Reference | Reference | Reference | Reference |
| Low Mental Wellbeing | **2.23 [1.17-4.22]** | **3.12 [1.54-6.32]** | 1.28 [0.73-2.23] | 2.05 [0.93-4.52] |

The socio-demographic observations discussed previously are also apparent in the multinomial logistic regression models. Females were two times (aOR 2.59 [95% CI 1.91-3.52]) and three times (aOR 3.39 [95% CI 2.22-5.16) more likely to experience moderate or high *wages* precariousness, respectively, compared to males (Table 6). The youngest age-group were almost two times (aOR 1.96 [95% CI 1.16-3.30) more likely to experience high *disempowerment* and three times (aOR 3.11 [95% CI 1.43-6.75]) more likely to experience high precariousness with regards to *rights*, compared to the 40-49 years age group (Tables 5 and 6). Moderate precariousness in relation to *rights* was experienced more across all age groups compared to 40-49 years and across all deprivation quintiles compared to the least deprived quintile (Table 6).

### Research Question IV. Has there been changes in job quality (as reflected by precarious employment domains) between February 2020 and Winter 2020/2021?

The employment trajectories of the 429 individuals (41.6% of baseline sample) who were in ‘paid employment’ both in February 2020 and the winter of 2020/2021 are presented in Figure 4. Of these, 88.6% were in the same job at both time points (Groups 1 and 3 in Figure 4).

**Figure 4.**
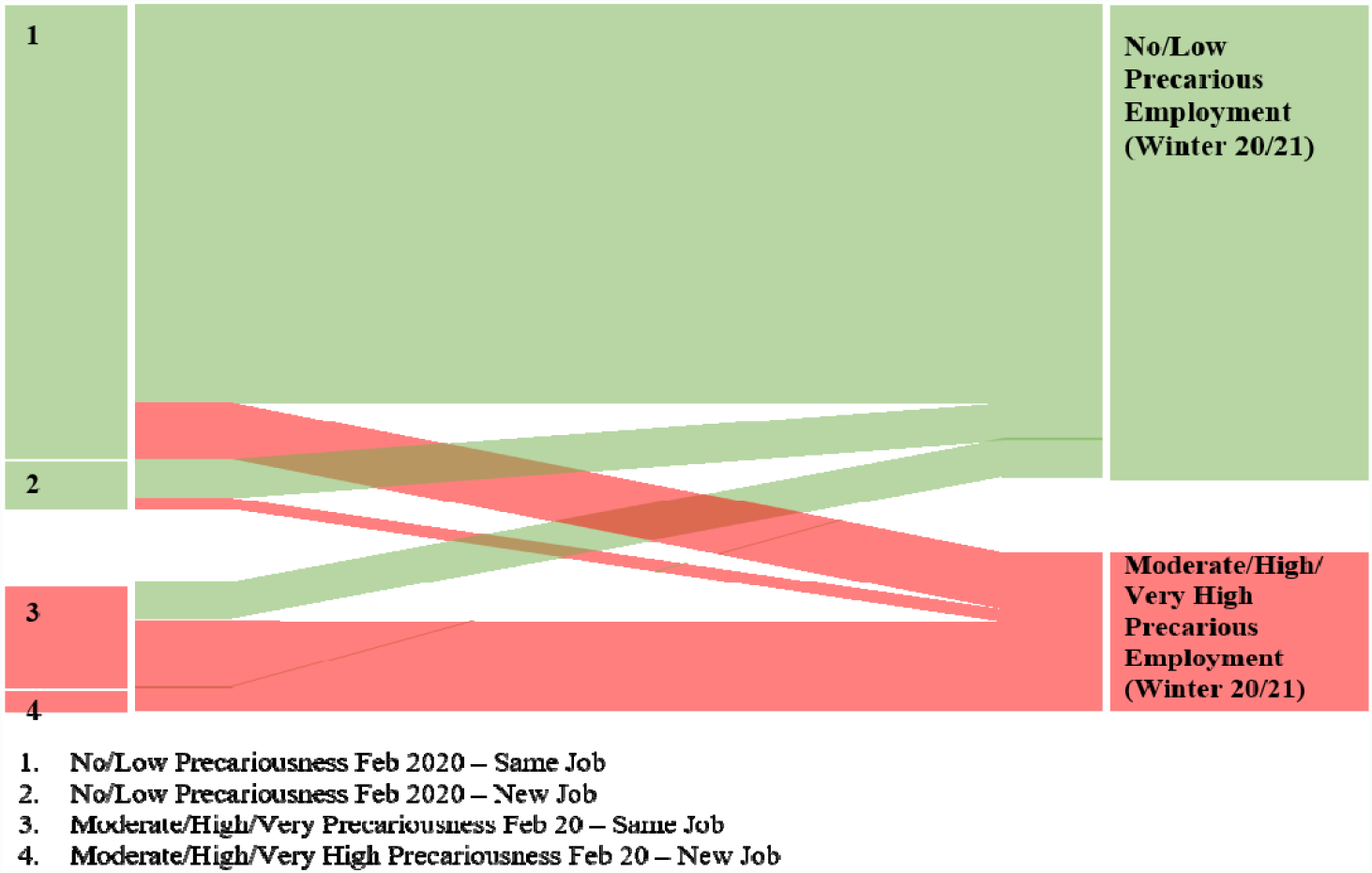
Trajectories of job quality (defined by precarious employment) experienced by the longitudinal sample since February 2020 (n=429).

Compared to February 2020, in Winter 2020/21 the proportion of individuals ‘in paid employment’ who met the definition of precarious employment had increased. In the follow-up sample, one in five jobs (20.0%) were determined to be moderately or higher precariousness at baseline (February 2020; Table 7) and this figure increased to one in four jobs (24.9%; p=0.019) in Winter 2020/21. This was largely due to people with no change in job describing aspects of their employment as more precarious (10.3%), and a smaller proportion changing job from those of no/low precariousness to those of moderate or higher precariousness. A small proportion of people with no change in job felt their employment was no longer of moderately or higher precariousness (6.6%), and an even smaller proportion moved from moderate or higher precariousness to jobs calculated to be of no/low precariousness (2.0%). Overall, 44.9% of individuals who have changed jobs since February 2020 are now employed in positions that are calculated as moderate or higher precariousness by the EPRES.

**Table 7.** Changes in prevalence of precarious employment from February 2020 (pre-COVID-19) to Winter 2020/2021 overall, and by socio-demographic variables (n=429).

|  | Precarious Employment (Feb 20) |  | Precarious Employment (Winter 20/21) |  |
| --- | --- | --- | --- | --- |
|  | No/Low | Moderate/High/Very High | No/Low | Moderate/High/Very High |
| All | 80.0% | 20.0% | 75.1%* | 24.9%* |
| *Gender* |  |  |  |  |
| Male | 81.5% | 18.5% | 79.5% | 20.5% |
| Female | 79.4% | 20.6% | 72.7%* | 27.3%* |
| p-value |  | 0.125 |  | **0.027** |
| *Age Group* |  |  |  |  |
| 18-29 Years | 74.4% | 25.6% | 69.2% | 30.8% |
| 30-39 Years | 82.4% | 17.6% | 73.6%* | 26.4%* |
| 40-49 Years | 79.0% | 21.0% | 74.8% | 25.2% |
| 50-59 Years | 83.2% | 16.8% | 80.0% | 20.0% |
| 60-64 Years | 74.1% | 25.9% | 70.4% | 29.6% |
| p-value |  | 0.532 |  | 0.546 |
| *Deprivation Quintile* |  |  |  |  |
| WIMD 1 | 81.1% | 18.9% | 75.7% | 24.3% |
| WIMD 2 | 72.5% | 27.5% | 68.8% | 31.2% |
| WIMD 3 | 81.4% | 18.6% | 70.0% | 30.0% |
| WIMD 4 | 79.7% | 20.3% | 77.2% | 22.8% |
| WIMD 5 | 86.6% | 13.4% | 83.5% | 16.5% |
| p-value |  | 0.157 |  | 0.129 |

Bold values denote significance between group analysis, \*denote significance within group analysis (P<0.05).

The drivers of this observed increase in precarious employment are likely attributable to an increased reported prevalence of higher wage precariousness in Winter 2020/21 (23.5% compared to 18.4%; p=0.009; Table 8) and more individuals reporting a sense of vulnerability at work (Table 8). There was a 6.3% and 3.0% increase in perceived moderate and high vulnerability, respectively, between February 2020 and Winter 2020/21 (Table 8). These changes cannot be explained alone by those who experienced furlough. In those who experienced furlough there was a 13.1% increase in reported moderate vulnerability (27.4% compared to 14.3%; p=0.027), however, those who did not experience furlough also reported increased vulnerability (Table 8). Both those who did and did not experience furlough reported an increase in high wages precariousness. Furthermore, the proportion of individuals holding the various contract types remained relatively unchanged, and some improvements in relation to access to rights were observed, especially in those who experienced furlough (+14.3% increase in low precariousness reported; Table 8). However, these improvements do not appear to offset the other domains which has resulted in an increase in precarious employment during the COVID-19 pandemic.

**Table 8.** Changes in precarious employment domains from February 2020 (pre-COVID-19) to Winter 2020/2021, overall and by experience of furlough during the pandemic.

|  | All<br>(n=429) |  |  | Did not<br>experience furlough<br>(n=344) |  |  | Experienced furlough<br>during pandemic<br>(n=84) |  |  |
| --- | --- | --- | --- | --- | --- | --- | --- | --- | --- |
|  | Feb<br>2020 | Winter<br>20/21 | %<br>Change | Feb<br>2020 | Winter<br>20/21 | %<br>Change | Feb<br>2020 | Winter<br>20/21 | %<br>Change |
| ***Temporariness (Contract Type)*** |  |  |  |  |  |  |  |  |  |
| Permanent | 90.9 | 90.0 | -0.9 | 91.6 | 91.9 | +0.3 | 89.3 | 83.4 | -5.9 |
| Fixed-Term | 5.4 | 7.0 | +1.6 | 5.8 | 6.4 | +0.6 | 2.4 | 8.3 | +5.9 |
| Atypical | 3.7 | 3.0 | -0.7 | 2.6 | 1.7 | -0.9 | 8.3 | 8.3 | 0.0 |
| ***Vulnerability*** |  |  |  |  |  |  |  |  |  |
| No/Low | 77.6 | 68.3 | -9.3* | 77.3 | 69.5 | -7.8* | 78.6 | 63.1 | -15.5* |
| Moderate | 18.2 | 24.5 | +6.3* | 19.2 | 23.8 | +4.6 | 14.3 | 27.4 | +13.1* |
| High/Very High | 4.2 | 7.2 | +3.0* | 3.5 | 6.7 | +3.2* | 7.1 | 9.5 | +2.4 |
| ***Disempowerment*** |  |  |  |  |  |  |  |  |  |
| No/Low | 49.2 | 52.4 | +3.2 | 48.5 | 52.9 | +4.4 | 52.4 | 51.2 | -1.2 |
| Moderate | 21.0 | 18.4 | -2.6 | 21.6 | 19.5 | -2.1 | 19.0 | 14.3 | -4.7 |
| High/Very High | 29.8 | 29.1 | -0.7 | 29.9 | 27.6 | -2.3 | 28.6 | 34.5 | +5.9 |
| ***Wages*** |  |  |  |  |  |  |  |  |  |
| No/Low | 42.0 | 38.9 | -3.1 | 46.5 | 43.6 | -2.9 | 23.8 | 20.2 | -3.6 |
| Moderate | 39.6 | 37.5 | -2.1 | 36.9 | 35.5 | -1.4 | 51.2 | 45.2 | -6.0 |
| High/Very High | 18.4 | 23.5 | +5.1* | 16.6 | 20.9 | +4.3* | 25.0 | 34.6 | +9.6 |
| ***Rights*** |  |  |  |  |  |  |  |  |  |
| No/Low | 72.5 | 79.3 | +6.8* | 78.2 | 83.1 | +4.9* | 48.8 | 63.1 | +14.3* |
| Moderate | 23.5 | 14.2 | -9.3* | 18.3 | 12.5 | -5.8* | 45.2 | 21.4 | -23.8* |
| High/Very High | 4.0 | 6.5 | +2.5 | 3.5 | 4.4 | +0.9 | 6.0 | 15.5 | +9.5* |

\*denote significance within group analysis (P<0.05).

## Discussion

Global evidence suggests that some population subgroups, namely younger people, migrant workers, and females are more likely to be in precarious employment [15–19]. Our data is consistent with this existing international evidence, and we observed precarious employment was more prevalent in females and in the youngest age group. Furthermore, we also observed that individuals who reported a pre-existing condition or low mental wellbeing were more likely to be in precarious employment than their healthier counterparts.

Our study shows that amongst respondents in February 2020, one in four (26.5%) of those in ‘paid employment’ in Wales were employed within roles that were precarious in nature at a moderate level or higher, as defined by five key domains (temporariness, disempowerment, vulnerability, wages and rights). This is despite the fact that 90% of all jobs were permanent contracts, highlighting the limitation of using contract type as a proxy for precarious employment [7,8]. European Data from 2014 calculated precarious employment in Anglo-Saxon countries (UK and Ireland) to be 58.5% [20]. Higher proportions were observed in Continental (69.3%), Eastern (72.6%) and Southern (67.0%) European regions, whereas Nordic countries (51.2%) had a lower prevalence of precarious employment [20]. However, the methodology to assess precarious employment in this study was different, with precarious employment determined if just one of four domains (temporariness; exercise rights; vulnerability; disempowerment) was experienced [20]. Our data shows that 84.8% of respondents experience at least one domain of precarious employment at a moderate level or higher in February 2020. With regards to individual domains of precarious employment, 25.4% of our respondents reported a sense of vulnerability, 50.8% reported disempowerment and 10.3% reported temporariness. In 2014 these values in UK and Ireland were 14.4% 27.7% and 11.0%, respectively [20]. Whilst the temporariness domain has remained relatively unchanged, proportions in the vulnerability and disempowerment domains were higher and may reflect the different context in 2020 given a global pandemic.

Through a combination of people reporting more precariousness in their job and individuals changing jobs to those calculated as precarious, there has been a resultant increase in the prevalence of precarious employment in Wales over the period from February 2020 to Winter 2020/21. This has been driven by increased proportions expressing increased vulnerability at work and an increased precariousness of *wages*. It is important to reiterate that these changes in relation to vulnerability were observed regardless of furlough status. These observations were also before the cost of living crisis, thus, it would be a fair assumption that precariousness of wages has worsened further, however follow-up analysis is required to confirm this. The negative impacts of the labour market changes through the pandemic have been experienced more by females than their male counterparts. Whilst the 18-29 years age group still have the highest proportion of individuals in precarious arrangements, there was a significant shift into precarious employment in those aged 30-39 years. Evidence demonstrates that those individuals holding non-permanent contract arrangements were more likely to have experienced unemployment in the early months (May/June 2020) of the pandemic [3]. Furthermore, since 1995, more than half of the new jobs created in the European Union have been part-time, non-contracted, or insecure positions [8,21]. Our data again follow this trend, with 44.9% of individuals who changed jobs during the pandemic moving into jobs calculated as precarious by the EPRES. Therefore, it is fair to assume that the quality of the labour market has become increasingly precarious in nature during the COVID-19 pandemic, somewhat providing confirmation of an informed prediction from the beginning of the pandemic that precarious employment would increase [6].

The cross-sectional and longitudinal observations both have population health implications. When examining the individual domains of precarious employment, it was vulnerability at work and low wages that were negatively associated with poorer health across all measures. This is a long-standing issue; as far back as 2005 the UK had a greater proportion of very low paid jobs compared to many other EU-27 countries [18]. The important relationship between better health and increased wealth has been established [22], however, it is also important to acknowledge the negative impacts of vulnerability and more specifically, perceived job insecurity at work on health. Perceived job insecurity is one prime example which can increase rates of sickness presenteeism rather than increase rates of sickness absence [23,24], and perceived job insecurity has been demonstrated to influence sickness absence in individuals who do not necessarily have contract insecurity [23]. Emerging research from the initial stages of the COVID-19 pandemic also reported associations between job insecurity, financial concerns and symptoms of anxiety [25].

Our data have also resurfaced two longstanding issues related to employment and health. One of which is low or insufficient pay in the UK, which is more prevalent than many other Western European nations [18]. The importance of a minimum income for healthy living has been proposed since the beginning of the century [26]. In practical terms, the provision of an adequate living wage in London resulted in better mental wellbeing in employees compared to those who do not receive the supplementary wage [27]. The second is that the creation of new jobs continues to be predominantly in non-permanent positions, thus removing the security and associated health benefits of good employment [28–30]. It is imperative that during the COVID-19 recovery phases the creation of secure, fairly paid employment opportunities are prioritised. If not, these longstanding issues will likely continue. Secondly, consistent with existing literature we observed a higher prevalence of precarious employment in females and those aged 18-29 years old [15–19]. These observations in isolation are therefore unsurprising. However, once the negative impacts of the COVID-19 pandemic on employment in the youngest age group [3,31] are considered alongside the well-established associations between job quality and health [28–30], there is a real concern for the long-term wellbeing of the future workforce in the UK and throughout Europe. Finally, the implications previously discussed are at an individual level, but there are also potential impacts on the wider community. The quality of the local labour market can result in reduced spending power and therefore a decline in community participation [8,32].

Despite the many strengths of this study including the first use of the EPRES in the UK, there are also a number of limitations that we wish to acknowledge. We recognise that this cross-sectional design cannot provide evidence of causality or direction of effect, therefore poorer health may be both a consequence and predictor of precarious employment. Further longitudinal analysis is required to investigate causality. Furthermore, within the context of a global pandemic we have to consider that there may be some response bias introduced into the data, especially on the sense of vulnerability and disempowerment. Also, due to inconsistencies in methodologies within the available international literature, direct comparisons between our data and previous figures (overall precariousness and by individual domains) should also be interpreted with some caution. In an attempt to address these methodology discrepancies, a 13 question Employment Precariousness Scale for Europe (EPRES-E) has been recently developed [33] (it should be noted that this scale was published after the initiation of our study). The authors concluded that the EPRES-E can be used for comparative purposes, although each individual domain score should also be reported [33]. Where possible, future research should be consistent with the approach used to determine precarious employment. Another important consideration to the longitudinal analysis is that those individuals who became unemployed or moved into self-employment were excluded from the analysis because the EPRES calculation is only applicable to ‘paid employment’.

## Conclusion

This study is the first to use a multi-dimensional tool to measure the prevalence of precarious employment in Wales. Our findings indicate that prior to the COVID-19 pandemic, one in four ‘paid employment’ jobs in Wales were calculated to be of moderate precariousness or higher as defined by the EPRES scale. The main domains contributing to employment precariousness in Wales were feelings of disempowerment (perceived lack of control of schedule) and wages which resulted in the inability to afford basic and unexpected expenses. The domains shown to be more closely related to poor self-reported health were insufficient wages to cover basic and/or unexpected expenses and vulnerability (defined as perceived unfair treatment, job insecurity or discrimination experienced at work). Improving these two domains, through the creation and provision of secure, adequately paid job opportunities has the potential to reduce the prevalence of precarious employment in Wales. In turn, these changes would improve health and wellbeing of the working age population, some of which are already adversely impacted by the COVID-19 pandemic. Further longitudinal studies would ascertain the directional impact of precarious employment, and inform the cross-government actions needed to address health and wellbeing in Wales.

## Data Availability

All relevant data produced in the present work are contained in the manuscript.

## Supplementary Data

### Precarious Employment Questionnaire

**Table S1.** Domains, questions and response options contained within the Employment Precariousness Scale (EPRES)

| **Domain** | **Questions** | **Responses (score)** |
| --- | --- | --- |
| *Temporariness (8)* | The duration of your contract is | Permanent contract (0)<br>1 year or more (1)<br>Temporary, non-fixed term (2)<br>Between 6 and less than 12 months (3)<br>Less than 6 months (4) |
|  | During the last twelve months, how long did you work under temporary contracts? | Did not work under temporary contracts (0)<br>For less than 2 months (1)<br>Between 2 and 3 months (2)<br>Between 3 and 6 months (3)<br>Between 6 and 12 months (4) |
| *Disempowerment (6)* | How do you settle the following employment conditions?<br>(a) Workplace schedule<br>(b) Weekly working hours<br>(c) Wages or salary | By collective agreement (0)<br>By the employer (1)<br>Don't know (2) |
| *Vulnerability (24)* | In relation to the way you are treated at work, can you tell me whether<br>(a) You feel afraid to demand better working conditions<br>(b) You feel defenceless towards unfair treatment by your superiors<br>(c) You feel afraid of being fired for not doing what you are asked to do<br>(d) You are treated in a discriminatory and unjust manner<br>(e) You are treated in an authoritarian and violent manner<br>(f) You are made to feel you can be easily replaced | Never (0)<br>Only one time (1)<br>Sometimes (2)<br>Many times (3)<br>Always (4) |
| *Wages (10)* | Does your current salary allow you to cover your basic needs? | Very much (0)<br>A good amount (1)<br>A little (2)<br>Not at all (3) |
|  | Does your current salary allow you to cover unexpected expenses? | Very much (0)<br>A good amount (1)<br>A little (2)<br>Not at all (3) |
|  | How much is your take home (net) monthly wage or salary? | More than €2406 (0)<br>€1504 to €2405 (1)<br>€752 to €1503 (2)<br>€452 to €751 (3)<br>€451 or less (4) |
| *Rights (14)* | Of the following benefits, which do you have a right to...<br>(a) Paid vacations<br>(b) Pension<br>(c) Severance pay<br>(d) Maternity/paternity leave<br>(e) Day off for family or personal reasons<br>(f) Weekly holidays<br>(g) Unemployment benefit/compensation | Yes (0)<br>No (1)<br>Don't know (2) |

